# Neurite Degradation Mediates the Effect of Amyloid Deposition on Global Cognition in Asymptomatic Older Adults

**DOI:** 10.1101/2025.07.23.25332087

**Authors:** Sasha Hakhu, Sydney Y. Schaefer, Scott C. Beeman, Alzheimer’s Disease Neuroimaging Initiative

## Abstract

This study investigates whether amyloid deposition is associated with neurite degradation in cognitively unimpaired (asymptomatic) older adults, with a focus on brain regions critical for memory and cognition. Using data from the ADNI3 cohort (n = 65; mean age 69.5±5.5), we examined the relationship between amyloid levels (from PET imaging), neurite density (derived from diffusion MRI-based Neurite Orientation Dispersion and Density Imaging, NODDI), and global cognition (MoCA scores). NODDI is a biophysical diffusion MRI model that quantifies microstructural features of brain tissue through metrics like neurite density index (NDI) that may sensitively capture early neurodegenerative changes leading up to Alzheimer’s Disease (AD). Spearman correlation analyses showed significant negative associations between amyloid and NDI in the right entorhinal cortex (ρ = -0.29; p = 0.02) and left fusiform gyrus (ρ = -0.26; p = 0.04). Amyloid also correlated with ODI in the left (ρ = -0.31; p = 0.011) and right fusiform gyrus (ρ = -0.33; p = 0.006). Mediation analyses revealed significant effects for NDI in the left entorhinal cortex (p = 0.02) and left fusiform gyrus (p = 0.006), and for ODI in the left fusiform gyrus (p = 0.044). These findings indicate that even in the absence of clinical symptoms, amyloid deposition may contribute to microstructural degradation in key brain areas, which in turn relates to cognitive function. NDI specifically may hold promise as an early imaging marker for identifying individuals at risk and tracking AD progression.

**Significance Statement:** This study demonstrates that neurite degradation, measured by neurite density index (NDI), mediates the relationship between amyloid deposition and global cognitive function in asymptomatic older adults, highlighting NDI as a promising early marker of microstructural vulnerability in preclinical Alzheimer’s disease.

## 1. Introduction

Alzheimer’s disease (AD) is characterized by early neuropathological changes, including amyloid-beta (Aβ) deposition, the emergence of tau tangles, and microstructural degeneration of neuronal tissue [1,2]. Amyloid positron emission tomography (PET) allows for the visualization of plaque accumulation within the brain, which is hypothesized to disrupt neuronal communication and trigger inflammation, leading to neurodegeneration prior to cognitive decline in AD patients [3,4]. While amyloid deposition has been extensively studied as a hallmark of AD [5,6], its impact on the integrity of the brain’s microstructure remains unclear, particularly among individuals who do not yet show cognitive symptoms (i.e., asymptomatic) and who may not yet show measurable volumetric loss as measured by standard structural neuroimaging methods (i.e., T1/T2-weighted magnetic resonance imaging). However, diffusion magnetic resonance imaging (dMRI) techniques like NODDI (Neurite Orientation Dispersion and Density Imaging) [7] allow for in vivo assessment of neurite microstructure [8,9]. NODDI is particularly advantageous for this purpose because it provides a detailed, quantitative measure of neurite density and orientation, which are crucial for assessing early neurodegenerative changes to which standard MRI methods are not sensitive. Such neurite degradation can be quantified through metrics such as neurite density index (NDI), orientation dispersion index (ODI), and isotropic volume fraction (fISO), which could aid in monitoring early changes in disease-specific brain regions prior to the onset of cognitive symptoms or volumetric loss [10,11]. These diffusion metrics derived via the NODDI model may therefore provide new insights within the Amyloid/Tau/Neurodegeneration (ATN) framework [12,13] that relate pathology and neurodegeneration in the context of dementia.

In support of this, associations between amyloid and neurite density have been documented in symptomatic patients diagnosed with AD [14–16], but it remains unclear whether this association is present in cognitively unimpaired older adults, and the extent to which this relationship leads to eventual cognitive impairment. Thus, this study aimed to investigate the association between PET-measured amyloid deposition and the neurite density index (NDI) in cognitively unimpaired individuals. This study focused primarily on the entorhinal cortex and fusiform gyrus, given that these regions have been shown to accumulate amyloid before substantial cognitive decline begins [17–20]. While regions like the primary motor cortex are less affected in very early stages of AD, they serve as useful controls to compare amyloid deposition and microstructural changes in unaffected areas [21–25]. We have shown previously that NDI in these regions differentiated cognitively-unimpaired APOE e4 carriers from non-carriers, whereas volumetric MRI did not [14]. Our prior study also showed a correlation between regional NDI and MoCA scores, whereas volumetric MRI did not, suggesting that NDI may capture clinically meaningful microstructural variation even in the preclinical phase. While we have evaluated NDI as well as ODI and fISO in asymptomatic older adults, NDI showed the strongest associations with key cognitive and genetic variables, and overall is most relevant to the current biological framework of Alzheimer’s disease since it quantifies neurite density—an aspect of microstructure that may decline before volumetric atrophy or cognitive impairment [12,13]. Thus, in the present study, we hypothesized that 1) higher amyloid levels would be associated with lower NDI in the entorhinal cortex and fusiform gyrus regions, and 2) NDI would mediate the relationship between amyloid levels and performance on the Montreal Cognitive Assessment (MoCA).

## 2. Methods

### 2.1. Participants

Data were obtained from the Alzheimer’s Disease Neuroimaging Initiative (ADNI3, 2017) database (adni.loni.usc.edu). This study analyzed data from 65 cognitively unimpaired older adults who underwent brain diffusion MRI, PET scanning and neuropsychological testing. The mean age of participants was 69.5 ± 5.5 years (range: 62–90), with 45 females.

### 2.2. Neuroimaging data acquisition

Data used in the preparation of this article were obtained from the Alzheimer’s Disease Neuroimaging Initiative (ADNI) database (adni.loni.usc.edu) [26]. The ADNI was launched in 2003 as a public-private partnership, with the primary goal of testing whether serial magnetic resonance imaging (MRI), positron emission tomography (PET), other biological markers, and clinical and neuropsychological assessment can be combined to measure the progression of mild cognitive impairment (MCI) and early Alzheimer’s disease (AD).

All MRI data were acquired using the ADNI-3 protocol on a 3T MRI scanner. The following sequences were used— (i) MPRAGE T1-weighted image (TR = 2300 ms, TI = 900 ms), (ii) multi-shell dMRI with three b-values (500, 1000, 2000 s/mm²), TR/TE = 3300/71 ms, with 112 diffusion-weighted directions and 13 interleaved b=0 images.

Amyloid pathology was assessed using PET imaging— 8F-Florbetapir (FBP) or 18F- Florbetaben (FBB) for Aβ pathology, depending on availability at the time of data collection. All PET data were preprocessed by the ADNI PET Core, following standardized protocols. Detailed acquisition and preprocessing methods are available on the ADNI website (https://adni.loni.usc.edu/wp-content/uploads/2012/10/ADNI3_PET-TechManual_V2.0_20161206.pdf).

### 2.3. Neuroimaging data processing

Preprocessing of diffusion data was conducted using FSL and MRtrix and included noise correction, susceptibility distortion correction (Synb0) [27], motion and eddy current correction [28], and bias field correction [29]. The diffusion model, NODDI (MATLAB toolbox) [30] was then applied to extract neurite density index (NDI), orientation dispersion index (ODI), and isotropic volume fraction (fISO) as diffusion metrics, all scaled from 0 to 1. As justified by our prior work [31], our primary diffusion metric was NDI, as it quantifies neurite volume fraction, with higher values indicating greater neurite density. Secondary diffusion metrics were also considered in this study; specifically, ODI measures the coherence of neurite orientation (where higher values reflect greater dispersion) and fISO represents the isotropic component of the diffusion signal (where higher values indicate more isotropic diffusion) that may be associated with increased water mobility in regions with less structured or aligned neural fibers. While all these metrics were computed and analyzed, the a priori hypothesis of this study is focused on NDI due to its stronger association with cognitive function and early vulnerability in AD-related brain regions [31].

T1 and b0 images underwent brain extraction and linear co-registration, followed by nonlinear transformation to the Montreal Neurological Institute (MNI) [32] standard brain space. Regions of interest (ROIs) were selected from the Harvard-Oxford cortical and subcortical structural atlases [33,34] and non-linearly mapped to an older brain atlas [35] to account for age-related structural changes, such as ventricular displacement. ROI-based mean diffusion metrics were extracted [36] for the fusiform gyrus and entorhinal cortex, based on prior research showing selective neurodegeneration in these regions within AD [17,18]. In addition, the left and right motor cortices were included as control regions to assess whether similar changes are present in areas typically spared during the early stages of the disease. Since previous studies have shown a stronger correlation between early dysfunction and volume loss in the left hemisphere compared to the right [37,38], we considered both the left and right ROIs.

### 2.4. Global cognition

Montreal Cognitive Assessment (MoCA) scores were analyzed to investigate associations between global cognition, amyloid deposition, and neurite microstructure. All participants were classified as cognitively unimpaired within the ADNI dataset based on extensive clinical evaluation, while the MoCA serves as a brief assessment of global cognition. Mean±SE scores on the MoCA in this sample were 25.75±0.34, ranging from 17 to 30 (maximum).

### 2.5. Statistical analysis

R software [39] was used to perform correlation analyses and were conducted for four key regions (left and right entorhinal cortex, and left and right fusiform gyrus) to assess the relationship between amyloid burden and neurite microstructure, while controlling for age and sex. Spearman’s rank correlation was used instead of Pearson’s correlation because normality assumptions were not met for several variables, i.e., Shapiro-Wilk tests were significant (p < 0.05) for amyloid and diffusion metrics. Spearman’s correlation coefficients, ρ values, and p-values were computed, with false discovery rate (FDR) corrections applied to adjust for multiple comparisons. Correlations were also run to test the relationship between regional amyloid deposition (SUVR) and cognitive performance (MoCA).

Mediation analysis was also performed using the “mediation” package in R to evaluate the role of diffusion metrics in linking regional amyloid burden (independent variable) to cognitive performance (MoCA, dependent variable). Our primary mediation analysis considered NDI as a mediator variable; however, additional separate analyses were run for fISO and ODI as mediator variables (see Supplementary Material). Linear models were fitted for both the mediator and outcome variables, controlling for age and sex. The mediation effects were quantified using the Average Causal Mediation Effect (ACME), which measured the indirect effect of amyloid on cognition through each diffusion metric. Statistical significance was determined using 95% confidence intervals and p-values derived from 1,000 bootstrap simulations.

## 3. Results

We computed correlation results between MoCA scores and regional amyloid deposition (measured as Standardized Uptake Value Ratio, or SUVR), as indicated by Spearman’s rank correlations (Supplementary Material Fig. S1 illustrates this monotonic relationship). Specifically, MoCA scores were significantly negatively correlated with amyloid deposition in the left entorhinal cortex (ρ = -0.3, p = 0.014), left fusiform gyrus (ρ = -0.27, p = 0.029), and left and right primary motor cortex (ρ = -0.30, p = 0.01 and ρ = -0.28, p = 0.024 respectively).

Next, Spearman’s rank correlation results between amyloid levels and diffusion metrics across entorhinal cortices, fusiform gyri, and primary motor cortices were computed. Significant negative correlations between amyloid deposition and NDI were observed in the right entorhinal cortex (ρ = -0.29, p = 0.018) and left fusiform gyrus (ρ = -0.26, p = 0.036), but not in their contralateral regions (p = 0.13 and p = 0.076, respectively). As hypothesized, amyloid was not significantly correlated with NDI in the left or right primary motor cortices (p = 0.97 and p = 0.72, respectively; Fig. S2). Amyloid levels were also negatively associated with ODI in both the left (ρ = -0.31, p = 0.011) and right fusiform gyrus (ρ = -0.33, p = 0.006), but not in the entorhinal cortex (all p > 0.5); and with fISO in the left motor cortex (ρ = 0.31, p=0.01). No significant correlations emerged between amyloid and fISO in any other region (all p > 0.1; Table S1).

Results from the mediation analysis assessing how NDI mediates the relationship between amyloid deposition and global cognition are shown in Table 1. NDI significantly mediated the relationship between amyloid levels and global cognitive function (MoCA) in the left entorhinal cortex (Average Causal Mediation Effect, ACME, p = 0.02) and left fusiform gyrus (ACME, p = 0.006), while mediation effects in the right hemisphere were not statistically significant (p > 0.05). However, even though the left and right primary motor cortices showed a significant relationship between amyloid level and MoCA, mediation analysis indicated that NDI was not a significant mediator of this relationship (all ACME p > 0.84). For the secondary diffusion metrics, a significant indirect effect was observed for ODI in the left fusiform gyrus (ACME, p = 0.044), whereas other ODI and fISO mediation effects were non-significant (all p > 0.14; Table S2). These overall findings suggest a causal link between amyloid and cognitive performance due to neurite degradation specifically within the entorhinal cortex and fusiform gyrus in asymptomatic individuals.

**Table 1.** Mediation analysis examining the indirect effects (average causal mediated effect (ACME)) of amyloid deposition on global cognition through neurite density index (NDI) in the entorhinal cortex, fusiform gyrus and motor cortex.

| **Region** | **Effect** | **Estimate** | **Lower**<br>**Confidence**<br>**Interval** | **Upper**<br>**Confidence**<br>**Interval** | **p** |
| --- | --- | --- | --- | --- | --- |
| **Left Entorhinal Cortex** | **ACME** | -2.43 | -5.41 | -0.21 | 0.02<sup>*</sup> |
| **Right Entorhinal Cortex** | **ACME** | -1.62 | -4.89 | 0.84 | 0.19 |
| **Left Fusiform Gyrus** | **ACME** | -2.27 | -4.60 | -0.50 | 0.006<sup>*</sup> |
| **Right Fusiform Gyrus** | **ACME** | -0.60 | -2.57 | 1.23 | 0.48 |
| **Left Motor Cortex** | **ACME** | -0.007 | -0.83 | 0.78 | 0.98 |
| **Right Motor Cortex** | **ACME** | 0.008 | -0.76 | 0.72 | 0.96 |

\*statistically significant

## 4. Discussion

The purpose of this study was to investigate the relationship between amyloid deposition and neurite density in cognitively unimpaired older adults. Our study expands on previous studies that demonstrate the association between amyloid deposition and neurite degradation in AD [40] by now demonstrating that amyloid deposition is associated with microstructural changes detected through diffusion magnetic resonance imaging (dMRI) that likely precede larger scale volumetric loss and cortical thinning as detected through standard structural neuroimaging methods (i.e., T1/T2-weighted magnetic resonance imaging). Results from this study showed negative associations between amyloid levels and neurite density in the entorhinal cortex and fusiform gyrus, aligning with the current framework of AD [13] suggesting that amyloid deposition disrupts neuronal integrity (measured as neurite density index, or NDI) in regions critical for memory and visuospatial cognition [18,41,42].

Importantly, these associations appear to be lateralized in our study, consistent with previous reports of asymmetrical vulnerability to neurodegeneration [37,43]. For instance, based on our results, although significant correlations between amyloid and both cognition and microstructural metrics were observed in both hemispheres, significant mediation effects emerged only in the left hemisphere. Specifically, NDI significantly mediated the relationship between amyloid and cognition in the left entorhinal cortex and left fusiform gyrus, while ODI showed a significant mediation effect only in the left fusiform gyrus. This lateralization aligns with prior studies reporting greater vulnerability of left hemispheric regions to early AD and may reflect the left hemisphere’s dominant role in memory, a cognitive domain highly sensitive to early AD-related decline [31,37,38,43,44]. Furthermore, the left entorhinal cortex and fusiform gyrus are closely linked to verbal memory and visuospatial processing [45–48], which are domains predominantly evaluated by the MoCA [49]. These findings underscore the importance of considering hemispheric asymmetries when identifying early Alzheimer’s disease markers.

A strength of this study is the inclusion of mediation analysis to test the extent to which NDI transmitted the effect of amyloid deposition on MoCA score (i.e., causal). Mediation analyses revealed that NDI significantly explained the relationship between amyloid and cognitive performance, particularly in the left entorhinal cortex and left fusiform gyrus. This suggests that amyloid-related microstructural degradation may be a key mechanism underlying early cognitive decline, even in individuals who are clinically unimpaired [50,51]. Although the strongest effects were observed in memory-related regions, we also extended these analyses to control regions like the left and right motor cortices to assess whether similar relationships emerged elsewhere. Here, neither did we observe correlations between amyloid and neurite density nor were there any significant mediation effects.

The fact that amyloid deposition was related to MoCA scores in the motor cortices but not mediated through NDI suggests that its cognitive effects in these regions may arise through non-microstructural mechanisms such as synaptic dysfunction, metabolic disruption, or large-scale network alterations. This aligns with evidence that amyloid-related cognitive decline can occur independently of detectable neurite loss in some areas [52–54]. Future studies will explore mechanisms by which motor cortical regions are associated with cognitive performance in older adults, particularly given that measures such as grip strength, gait speed, and finger tapping have been associated with cognitive impairment but may not necessarily be specific to AD [55–57]. At the same time, the results highlight the sensitivity of NDI to amyloid-related structural degeneration that may contribute to cognitive decline, particularly in regions that are early targets of AD.

Results from this study also suggest that ODI and fISO (other dMRI metrics) may be less valuable as early indicators of amyloid-driven neurodegeneration. No significant mediated effects were observed for fISO in any region, nor for ODI outside these left-lateralized region. NDI, however, showed region-specific evidence in capturing amyloid-related microstructural contributions to cognitive performance in older adults, consistent with our primary hypothesis. Thus, this study further reinforces the idea that different microstructural markers may capture distinct aspects of amyloid-related brain changes [58–60].

Several limitations should be considered. First, this study was cross-sectional and only evaluated mediation between regional amyloid and cognitive performance at a single timepoint. Longitudinal studies are necessary to better establish causal relationships and track the progression of these changes over time [61]. Future studies with larger cohorts and imaging techniques, such as the implementation of more complex diffusion microstructure models like Soma and Neurite Density Imaging (SANDI) [62,63] could provide a more comprehensive understanding of neurite integrity and how changes as over time with the accumulation of AD-specific neuropathology (amyloid and tau). Second, this study did not evaluate tau-PET, given that tau burden is more strongly linked to neurodegeneration and cognitive impairment in symptomatic stages of AD [64–66]. Lastly, cognitive performance was assessed using a global measure (MoCA), which may not capture subtle domain-specific deficits that are evaluated through an extensive neuropsychological battery. In short, however, this study provides important preliminary evidence of a relationship between amyloid deposition and neurite density in cognitively unimpaired individuals, suggesting that early changes in brain microstructure may occur before the onset of cognitive symptoms.

## Data Availability

Alzheimers Disease Neuroimaging Initiative publicly available data were used in this study.

https://adni.loni.usc.edu/

## Acknowledgments

Data collection and sharing for this project was funded by the Alzheimer’s Disease Neuroimaging Initiative (ADNI) (National Institutes of Health Grant U01 AG024904) and DOD ADNI (Department of Defense award number W81XWH-12-2-0012). ADNI is funded by the National Institute on Aging, the National Institute of Biomedical Imaging and Bioengineering, and through generous contributions from the following: AbbVie, Alzheimer’s Association; Alzheimer’s Drug Discovery Foundation; Araclon Biotech; BioClinica, Inc.; Biogen; Bristol-Myers Squibb Company; CereSpir, Inc.; Cogstate; Eisai Inc.; Elan Pharmaceuticals, Inc.; Eli Lilly and Company; EuroImmun; F. Hoffmann-La Roche Ltd and its affiliated company Genentech, Inc.; Fujirebio; GE Healthcare; IXICO Ltd.;Janssen Alzheimer Immunotherapy Research & Development, LLC.; Johnson & Johnson Pharmaceutical Research & Development LLC.; Lumosity; Lundbeck; Merck & Co., Inc.;Meso Scale Diagnostics, LLC.; NeuroRx Research; Neurotrack Technologies; Novartis Pharmaceuticals Corporation; Pfizer Inc.; Piramal Imaging; Servier; Takeda Pharmaceutical Company; and Transition Therapeutics. The Canadian Institutes of Health Research is providing funds to support ADNI clinical sites in Canada. Private sector contributions are facilitated by the Foundation for the National Institutes of Health (^36,37^). The grantee organization is the Northern California Institute for Research and Education, and the study is coordinated by the Alzheimer’s Therapeutic Research Institute at the University of Southern California. ADNI data are disseminated by the Laboratory for Neuro Imaging at the University of Southern California.

Dr. Sydney Schaefer is affiliated with the Arizona Alzheimer’s Consortium, and is a REC Fellow of the Arizona Alzheimer’s Disease Research Center (P30 AG072980).

## Consent Statement

All human subjects participating in studies conducted by the Alzheimer’s Disease Neuroimaging Initiative (ADNI), provided informed consent.

## Supplemntary Figures

**Figure S1.**
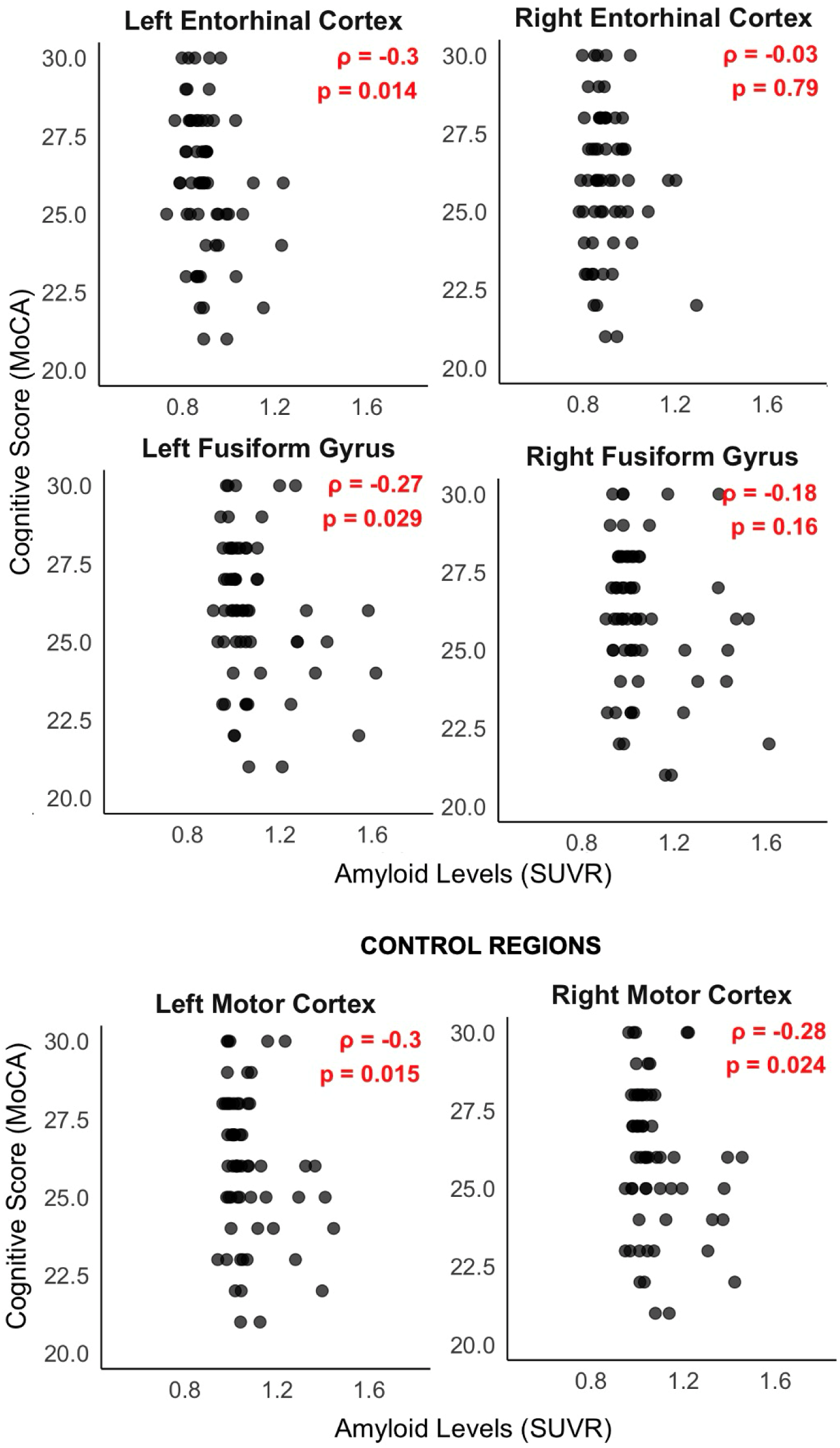
Spearman rank correlations between amyloid levels (SUVR (Standardized Uptake Value Ratio)) and global cognition (MoCA scores) in left and right— entorhinal cortices, fusiform gyri and motor cortices.

**Figure S2.**
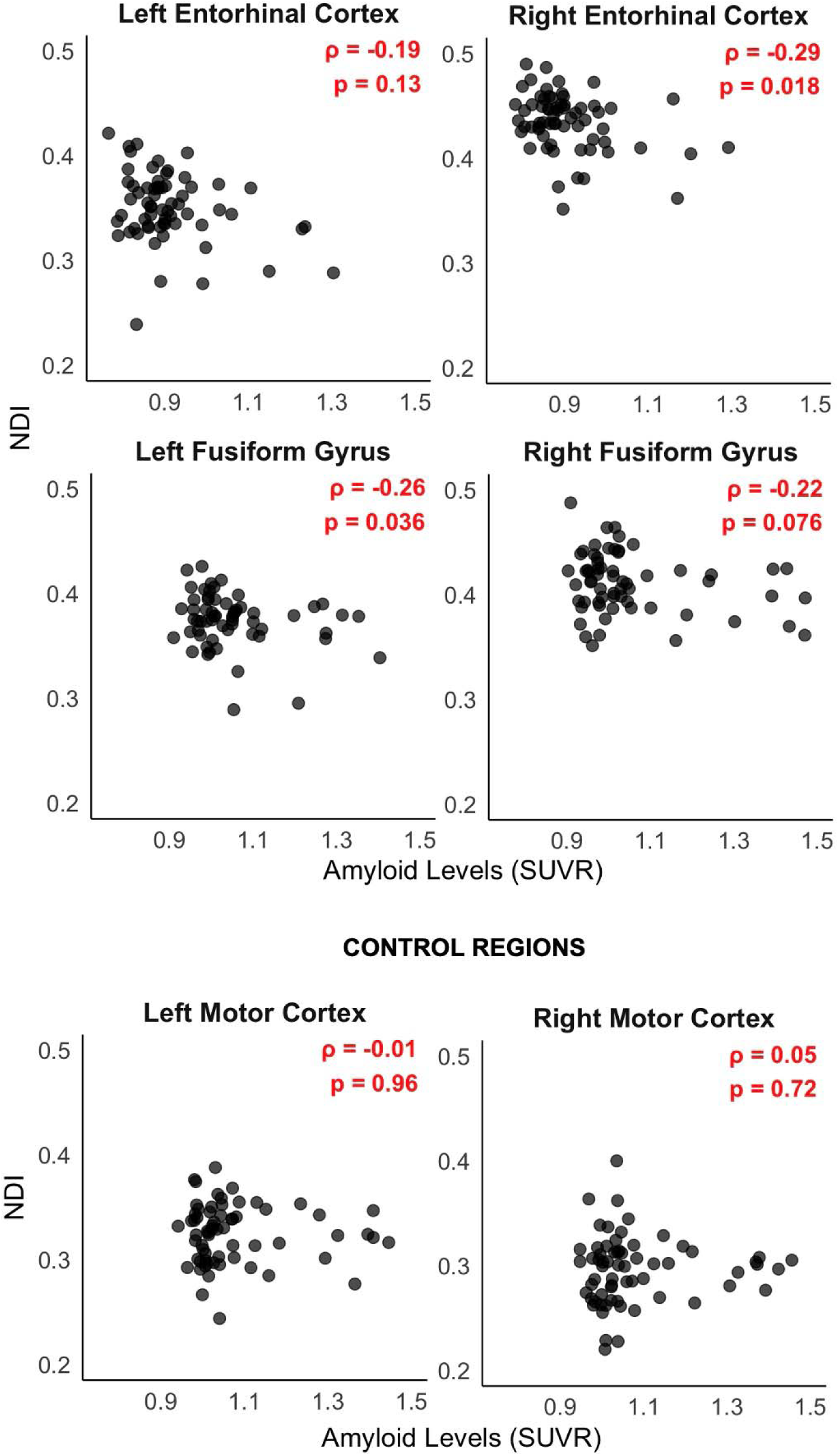
Spearman rank correlations between amyloid levels (SUVR (Standardized Uptake Value Ratio)) and neurite density index (NDI) in the left and right— entorhinal cortices, fusiform gyri and motor cortices.

## Supplementary Tables

**Table S1.** Correlation analysis examining associations between amyloid deposition and diffusion metrics— orientation dispersion index (ODI) and isotropic volume fraction (fISO) in the entorhinal cortex, fusiform gyrus and motor cortex.

| **Amyloid-ODI** |  |  |
| --- | --- | --- |
| **Region** | **$\rho$** | **p** |
| **Left Entorhinal Cortex** | -0.05 | 0.69 |
| **Right Entorhinal Cortex** | -0.07 | 0.58 |
| **Left Fusiform Gyrus** | -0.31 | 0.01* |
| **Right Fusiform Gyrus** | -0.33 | 0.007* |
| **Left Motor Cortex** | 0.09 | 0.46 |
| **Right Motor Cortex** | 0.14 | 0.28 |
| **Amyloid-fISO** |  |  |
| **Region** | **$\rho$** | **p** |
| **Left Entorhinal Cortex** | -0.13 | 0.71 |
| **Right Entorhinal Cortex** | -0.32 | 0.44 |
| **Left Fusiform Gyrus** | -0.21 | 0.64 |
| **Right Fusiform Gyrus** | 0.08 | 0.71 |
| **Left Motor Cortex** | 0.31 | 0.01* |
| **Right Motor Cortex** | 0.2 | 0.11 |

\*statistically significant

**Table S2.** Mediation analysis examining the indirect effects (average causal mediated effect (ACME)) of amyloid deposition on global cognition through diffusion metrics— orientation dispersion index (ODI) and isotropic volume fraction (fISO) in the entorhinal cortex, fusiform gyrus and motor cortex.

| **MoCA-Amyloid-ODI** |  |  |  |  |  |
| --- | --- | --- | --- | --- | --- |
| **Region** | **Effect** | **Estimate** | **Lower**<br>**Confidence**<br>**Interval** | **Upper**<br>**Confidence**<br>**Interval** | **p** |
| **Left Entorhinal Cortex** | **ACME** | -1.21 | -3.57 | 0.46 | 0.15 |
| **Right Entorhinal Cortex** | **ACME** | -0.80 | -3.28 | 1.36 | 0.47 |
| **Left Fusiform Gyrus** | **ACME** | -1.48 | -3.44 | -0.03 | 0.044* |
| **Right Fusiform Gyrus** | **ACME** | -0.11 | -1.64 | 1.48 | 0.88 |
| **Left Motor Cortex** | **ACME** | 0.007 | -0.82 | 0.82 | 0.98 |
| **Right Motor Cortex** | **ACME** | 0.004 | -0.91 | 0.98 | 0.99 |
| **MoCA-Amyloid-fISO** |  |  |  |  |  |
| **Region** | **Effect** | **Estimate** | **Lower**<br>**Confidence**<br>**Interval** | **Upper**<br>**Confidence**<br>**Interval** | **p** |
| **Left Entorhinal Cortex** | **ACME** | -0.79 | -2.98 | 0.98 | 0.38 |
| **Right Entorhinal Cortex** | **ACME** | -0.12 | -3.05 | 2.74 | 0.94 |
| **Left Fusiform Gyrus** | **ACME** | -0.68 | -2.58 | 0.91 | 0.37 |
| **Right Fusiform Gyrus** | **ACME** | -0.19 | -1.53 | 0.96 | 0.71 |
| **Left Motor Cortex** | **ACME** | 0.36 | -1.22 | 2.18 | 0.64 |
| **Right Motor Cortex** | **ACME** | 0.56 | -1.00 | 2.48 | 0.46 |

\*statistically significant

